# Runtime Anomaly Detection and Assurance Framework for AI-Driven Nurse Call Systems

**DOI:** 10.64898/2026.03.16.26348563

**Authors:** Yuanyuan Liu, David R. Concepcion

## Abstract

This research proposes an anomaly detection and assurance framework. It is mainly aimed at providing a framework for anomaly detection and assurance in AI-driven Nurse Call Systems (NCS) during operation. This study detects abnormal behaviors through simulating real call logs, injecting controllable anomalies, and using a lightweight Isolation Forest model. The final visualization results are presented through an interactive dashboard. Our research focuses mainly on the medical environment, which has characteristics of being delay-sensitive and safety-critical. A distinctive feature of this research is that it can effectively enhance the reliability of system operation without relying on complex deep model proprietary data, while maintaining safety and interpretability. The framework design emphasizes reproducibility while maintaining low computational overhead. The purpose is to enable rapid deployment of this framework on resource-constrained edge devices. Preliminary experimental results show that this method can maintain a reasonable precision rate. Additionally, when detecting delay-type anomalies, the results indicate a high recall rate. Moreover, to reflect the system’s performance in real scenarios, the framework detects delay metrics and hourly alarm quantity metrics, and reports Precision-Recall curves and their confidence intervals. Future work will consider introducing time, context features, and explainability analysis modules. The aim is to improve the model’s accuracy and further meet the medical industry’s requirements for auditability. This work focuses on the operational safety and reliability of AI-enabled Nurse Call Systems, addressing runtime failure modes that are underrepresented in current healthcare AI deployments. Rather than proposing new learning models, the contribution lies in a reproducible, interpretable assurance framework suitable for real clinical infrastructure. To ensure transparency and reproducibility, all code, cleaned datasets, experiment scripts, and an interactive Streamlit demo—allowing users to upload their own CSVs — are publicly released as open research artifacts (Zenodo DOI: 10.5281/zenodo.17767143).

## I. Introduction

Nurse Call Systems (NCS) are a critical component of hospital infrastructure, serving as the primary interface through which patients request assistance and clinical staff coordinate timely responses. In safety-critical care environments, delays, missed calls, or repeated false alarms can directly affect patient outcomes and place additional cognitive and operational burden on clinical teams. As hospitals scale and workloads fluctuate across time and units, the reliability of these systems becomes increasingly important for maintaining patient safety and care quality.

Recent deployments of artificial intelligence and Internet of Things technologies have enhanced the functionality of modern NCS, enabling features such as automated prioritization, workload balancing, and predictive monitoring. However, while these systems emphasize improved efficiency, there is comparatively less attention given to runtime assurance of learning-enabled components once deployed. In practice, abnormal behaviors such as excessive response delays, repeated alerts, or silent failures may not be detected in real time, leaving clinical staff unaware of emerging system risks. This gap highlights the need for lightweight mechanisms that can monitor operational behavior continuously and surface safety-relevant anomalies during live operation.

This work addresses that gap by proposing a reproducible runtime anomaly detection and assurance framework tailored to AI-enabled Nurse Call Systems. Rather than introducing new learning algorithms, the contribution lies in applying interpretable, lightweight anomaly detection techniques within an operational monitoring pipeline that reflects real clinical constraints. The framework emphasizes transparency, low computational overhead, and deployability on resource-constrained infrastructure, while providing actionable indicators of abnormal system behavior. Through controlled simulation and evaluation, the approach demonstrates how runtime monitoring can enhance the safety and reliability of AI-assisted clinical systems.

To operationalize this framework, the study evaluates runtime behavior using simulated nurse call logs with injected, clinically motivated anomalies, enabling controlled analysis of delay patterns, repeated alerts, and missing responses. A lightweight unsupervised anomaly detection baseline is employed alongside a simple rule-based comparator to assess detection performance under realistic workload conditions. Evaluation focuses on operationally meaningful metrics, including detection accuracy, response delay sensitivity, and alarm frequency, with results presented through an interactive visualization layer to support interpretability and auditability.

## II. Problem Definition

Nurse Call Systems (NCS) are safety-critical components of hospital operations, responsible for mediating timely communication between patients and clinical staff. In environments where workload, staffing, and patient acuity fluctuate continuously, abnormal system behavior such as prolonged response delays, repeated alerts, missing responses, or unexpected surges in call volume can introduce significant patient safety risks. These behaviors may arise from operational stress, device faults, network disruptions, or downstream effects of learning-enabled components integrated into the system.

As modern NCS increasingly incorporate artificial intelligence to support prioritization, routing, and workload management, a key challenge emerges after deployment: ensuring that learning-enabled components continue to behave reliably during live operation. While existing systems emphasize functional capability and efficiency, they typically provide limited mechanisms for continuously monitoring runtime behavior or detecting safety-relevant deviations as they occur. As a result, abnormal conditions may persist unnoticed until they escalate into clinical incidents.

Traditional approaches to assurance often depend on complex models, large labeled datasets, or offline validation, making them difficult to apply in real-time, resource-constrained clinical environments. In contrast, healthcare infrastructure requires assurance mechanisms that are lightweight, interpretable, and capable of operating continuously with low latency. This mismatch creates a gap between the growing use of AI-enabled functionality in nurse call systems and the ability to provide ongoing operational assurance once those systems are deployed.

Accordingly, the research question addressed in this work is: How can lightweight runtime monitoring and anomaly detection be used to improve the safety and reliability of AI-enabled Nurse Call Systems during live operation? The central hypothesis is that integrating simple, interpretable anomaly detection techniques into a runtime assurance framework can surface clinically meaningful deviations such as excessive response delays or abnormal call patterns, without imposing significant computational burden or relying on proprietary clinical data.

These requirements motivate the design of a continuous runtime monitoring approach that prioritizes deployability, transparency, and operational relevance over algorithmic complexity.

## III. Related Work

Prior research on anomaly detection has primarily focused on improving accuracy, latency, and reliability, particularly within medical and Internet of Things (IoT) systems. For example, Said et al. [1] proposed an anomaly detection framework for hospital IoT environments based on edge computing, while Tabassum et al. [2] introduced a multimodal hybrid approach to address sensor noise in smart healthcare settings. Related work has also demonstrated the feasibility of real-time anomaly detection in mission-critical communication systems [3], and indirect or weakly supervised learning approaches have been explored to mitigate the challenge of limited medical event annotation [4].

More recently, explainability and governance have emerged as central considerations for the credibility and adoption of medical artificial intelligence. Amann et al. [5] and Sadeghi et al. [6] provided systematic reviews of the technical, ethical, and regulatory roles of explainability in healthcare AI. Specific explainability techniques for tabular and time-series data have been discussed by Di Martino and Delmastro [7], and the relationship between explainability and security assurance has been explicitly examined by Jia et al. [8]. At the organizational level, Kim et al. and Rees et al. [9], along with policy-oriented work from Duke Health [10], emphasized the importance of governance frameworks for managing risk and accountability in clinical AI deployments. Information governance has further been framed as a socio-technical process influencing system trust and oversight [11].

Taken together, these studies demonstrate the feasibility of applying lightweight machine learning techniques, including unsupervised methods, to anomaly detection in healthcare-related systems. They also highlight the growing importance of interpretability and governance as prerequisites for deploying AI in safety-critical environments. However, much of the existing literature concentrates on general IoT platforms, communication systems, or offline evaluation settings, with comparatively limited attention to Nurse Call Systems as a distinct clinical infrastructure.

In particular, there remains a gap in research that integrates anomaly detection with continuous runtime assurance tailored to nurse call operations, where response delays and alert prioritization have direct safety implications. While prior work establishes relevant anomaly types, performance metrics, and interpretability techniques, validation in the context of real-time nurse call prioritization and operational monitoring remains limited. This gap motivates the framework proposed in this work, which focuses on runtime behavior, deployability, and assurance in AI-enabled nurse call systems.

In response to the gaps identified above, the following section presents a lightweight runtime assurance framework tailored to AI-enabled nurse call systems.

## IV. System Proposal

This work proposes a lightweight runtime anomaly detection and assurance framework designed to monitor the operational behavior of AI-enabled Nurse Call Systems (NCS) during live deployment. The framework directly addresses the assurance requirements identified in the preceding section by focusing on safety-relevant deviations such as excessive response delays, repeated calls, missing responses, and abnormal call volumes, while maintaining low computational overhead and high interpretability.

The framework employs a lightweight unsupervised anomaly detection approach selected for interpretability, computational efficiency, and suitability for continuous runtime monitoring in clinical environments. Rather than relying on complex deep learning architectures or large labeled datasets, the approach detects deviations from normal operational patterns using features derived from nurse call logs. Within this framework, Isolation Forest is used as the primary unsupervised baseline model due to its efficiency in identifying outliers and its compatibility with real-time deployment constraints.

To assess whether learning-based detection provides operational benefit beyond simple heuristics, the framework incorporates a rule-based baseline, such as flagging calls with response delays exceeding a predefined threshold. This comparison emphasizes practical runtime performance rather than algorithmic novelty, reflecting how such systems would be evaluated in real clinical settings.

The overall system architecture consists of four modular components:

- **Log generator:** simulates structured nurse call events, including timestamps and response delays;
- **Anomaly injector:** introduces clinically motivated abnormal behaviors into the logs;
- **Anomaly detector:** analyzes runtime patterns and flags potential anomalies;
- **Result visualizer:** presents results through an interactive dashboard

**Fig. 1.**
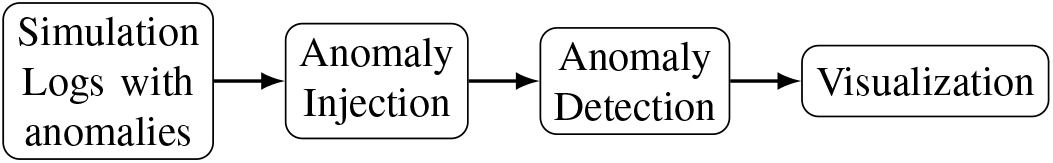
Overall architecture — Simulation → Anomaly Injection → Anomaly Detection → Visualization.

This modular design supports extensibility, allowing future integration of temporal context features, additional anomaly types, or explainability techniques without restructuring the core pipeline.

By combining lightweight anomaly detection with runtime monitoring and visualization, the proposed system provides a practical assurance mechanism that enhances transparency and situational awareness in AI-assisted nurse call operations. The framework is intended to complement, rather than replace, existing clinical workflows, offering an additional layer of safety oversight for learning-enabled components deployed in healthcare infrastructure.

## V. Experimental Setup

### A. Data Generation and Anomaly Injection

To evaluate the proposed runtime assurance framework under controlled yet operationally realistic conditions, the experimental setup uses simulated nurse call logs that reflect typical hospital communication patterns. Each log entry includes structured attributes such as room identifier, timestamp, call type, and response delay. Simulation enables systematic control over workload characteristics and anomaly behavior while avoiding reliance on proprietary clinical data.

Clinically motivated anomalies are injected into the simulated logs to represent safety-relevant deviations observed in real-world nurse call operations. These include excessive response delays, repeated calls from the same location within short time intervals, missing responses, inconsistent timestamps, and abnormal bursts in call volume. Table I summarizes the mapping between injected anomaly behaviors and their corresponding real-world interpretations. This design ensures that evaluation focuses on deviations with clear operational and clinical significance.

### B. Models and Baselines

The anomaly detection component is evaluated using a lightweight unsupervised approach consistent with the deployment constraints identified earlier. Isolation Forest is selected as the primary unsupervised baseline due to its computational efficiency, scalability, and suitability for detecting outliers in tabular operational data. Additional models are included to contextualize performance across different detection paradigms.

To assess whether learning-based detection provides benefit beyond simple heuristics, a rule-based baseline is implemented. This baseline flags anomalies when response delays exceed a predefined threshold, reflecting common operational rules used in practice. The inclusion of this baseline enables evaluation of whether the proposed approach offers meaningful improvement over fixed, manually defined criteria, emphasizing practical runtime performance rather than algorithmic novelty.

### C. Experimental Design and Procedure

The experimental design varies several **independent variables**, including injected anomaly rate, response delay thresholds, workload intensity, and the anomaly detection method employed. These variables are systematically adjusted to evaluate model behavior under different operational conditions.

Each experimental configuration is executed across multiple independent runs to account for stochastic variation in data generation and model behavior. Unless otherwise stated, results are averaged over ten runs. Ground truth labels are known due to the controlled anomaly injection process, enabling precise measurement of detection performance. This evaluation strategy emphasizes runtime realism, focusing on how models behave under varying load conditions rather than optimizing for offline accuracy alone.

### D. Evaluation Metrics and Operational Criteria

The **dependent variables** include both classification-based and operational metrics. Classification performance is measured using Precision, Recall, and F1-score, along with Precision–Recall (PR) curves, which are more informative than Receiver Operating Characteristic (ROC) curves under the class imbalance typical of safety-critical alerting systems. Recall is emphasized due to its relevance for patient safety, as missed anomalies may correspond to delayed or unaddressed clinical events. F1-score is used to assess the balance between detection sensitivity and false alarm burden.

In addition to classification metrics, operational metrics are included to reflect deployment constraints, such as detection delay and alarm frequency per hour. These measures capture the trade-offs between timely detection and alert fatigue, which are central considerations in clinical environments.

### E. Threshold Sensitivity Analysis

Rather than fixing alert thresholds a priori, the framework performs threshold sensitivity analysis by sweeping anomaly score thresholds across a defined range. This process identifies operating points that balance detection performance and operational burden, enabling evidence-based configuration of alert thresholds. Threshold values are therefore treated as tunable parameters derived from evaluation outcomes, rather than as design assumptions.

-Anomaly Mapping Table: Mapping of Simulated Anomalies to Real-World Scenarios

This table shows how each simulated anomaly corresponds to a specific injection method and its real-world interpretation in nurse call systems

Table I summarizes the mapping between injected anomaly behaviors and their corresponding operational interpretations in nurse call systems. These anomalies are designed to represent safety-relevant deviations that may occur during live operation, rather than to model specific clinical diagnoses or root causes. This abstraction enables systematic evaluation of runtime monitoring performance while preserving clinical plausibility.

**TABLE 1.** Mapping of Injected Anomalies to Operational Interpretations in Nurse Call Systems.

| Anomaly Type | Injected Behavior | Operational Interpretation |
| --- | --- | --- |
| Delayed Response | Response delay exceeding 90 seconds | Staffing overload, routing delay, or workflow disruption |
| Repeated Call | Multiple calls from the same room within 30 seconds | Patient distress, unacknowledged call, or system double-trigger |
| Missing Response | Removal of an expected log entry | System dropout, communication failure, or data loss |
| Invalid Timestamp | Event timestamps inconsistent with call order | Clock desynchronization or device fault |
| Unusual Call Volume | Burst of 10 or more calls within one minute | Emergency condition or malfunctioning sensor |

## VI. Experimental Results

This section presents the experimental results evaluating the proposed runtime anomaly detection and assurance framework for nurse call systems. The results focus on operational behavior, detection performance, and interpretability, with the goal of assessing whether lightweight anomaly detection can support continuous runtime assurance in safety-critical clinical environments.

### A. Workload Characteristics and Operational Patterns

To characterize baseline system behavior, simulated nurse call logs were generated using cleaned NYC 311 service request data, resulting in more than ten thousand records. Analysis of response times reveals a long-tailed distribution with clear daily and weekly periodicity, as illustrated in Figure 2. These temporal patterns indicate predictable workload fluctuations, supporting the suitability of continuous runtime monitoring for detecting deviations from expected operational behavior.

**Fig. 2.**
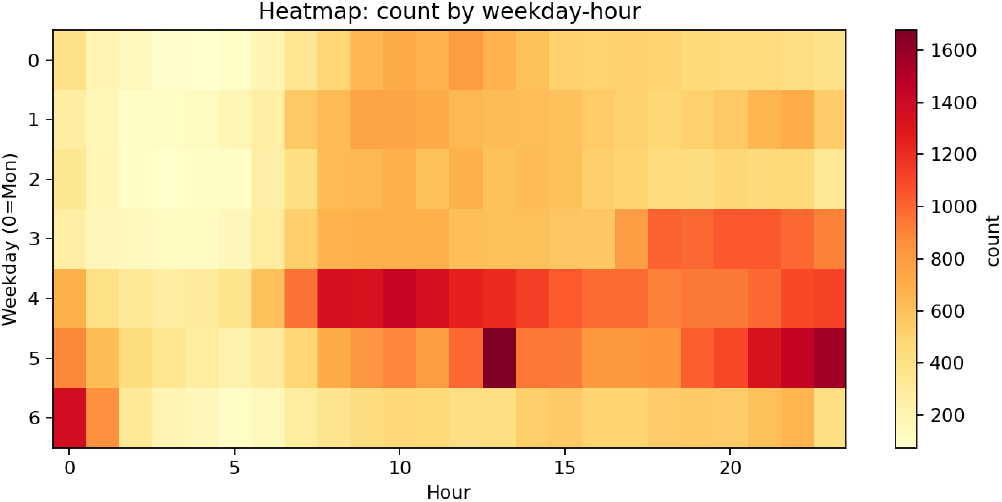
Heatmap by weekday-hour — workload periodicity across time.

### B. Detection Performance Across Models

Figure 3 summarizes detection performance across evaluated models using mean metrics with 95% confidence intervals. Among the supervised baselines, Random Forest achieved the highest overall performance (Precision ≈1.00, Recall ≈0.98, F1≈ 0.97), reflecting its effectiveness when labeled data are available. However, reliance on supervised training limits its applicability in many real-world deployment scenarios.

**Fig. 3.**
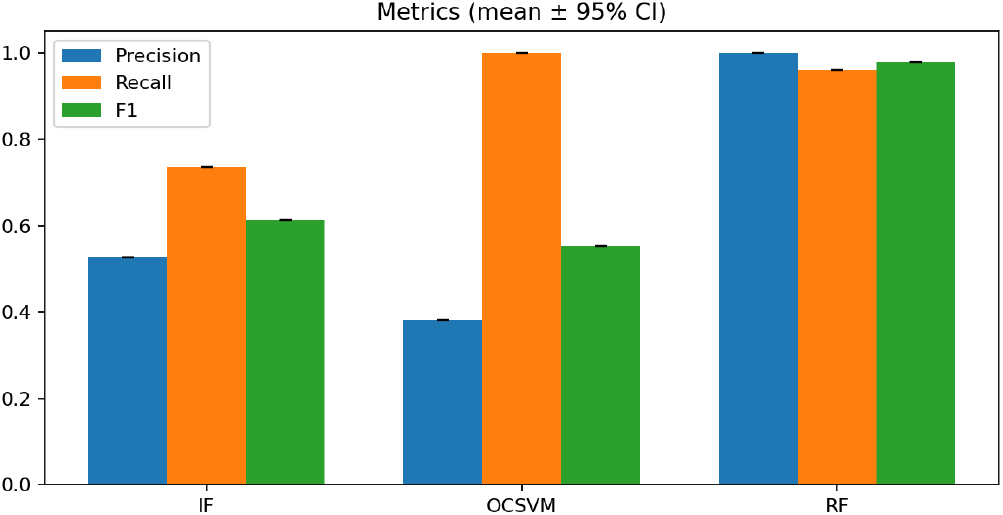
Metrics comparison (mean ±95% CI) — quantitative performance summary.

Isolation Forest demonstrated robuswell-suitedce under unsupervised conditions, with stable behavior across threshold settings and favorable trade-offs between detection sensitivity and false alarm rate. These characteristics make it well suited for real-time deployment where labeled data are limited and computational efficiency is required. In contrast, One-Class SVM exhibited sensitivity to noise and lower overall accuracy, while the Autoencoder achieved moderate performance (average precision ≈0.42), indicating potential supplementary value but higher complexity and tuning overhead.

Precision–Recall curves for the primary models are shown in Figure 4. As expected under class imbalance, PR curves provide a more informative view of detection performance than Receiver Operating Characteristic (ROC) curves. Isolation Forest maintains a favorable balance between precision and recall across operating points, supporting its use within a runtime assurance context.

**Fig. 4.**
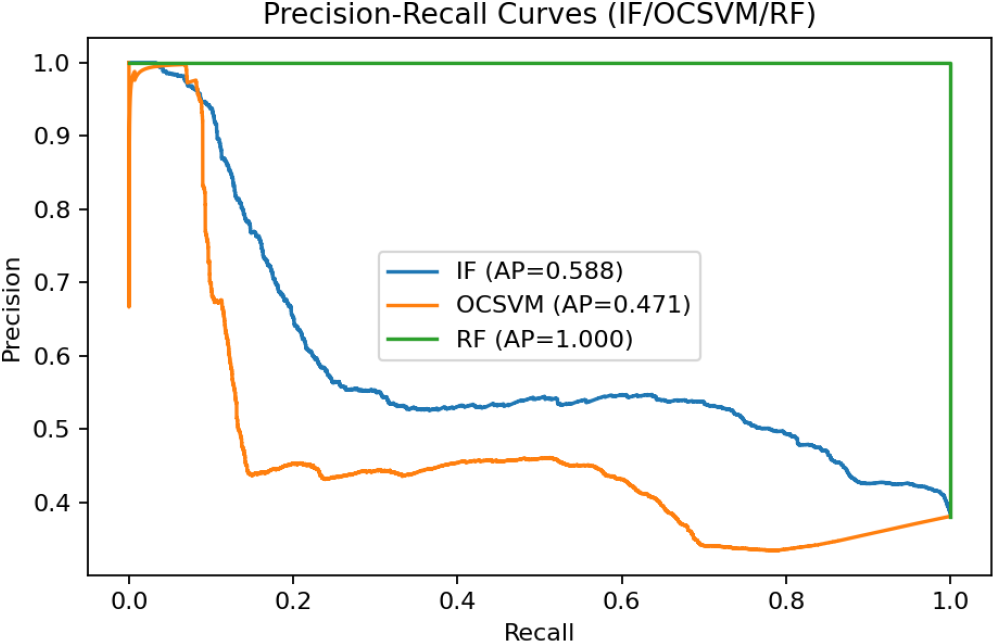
Precision–Recall curves (IF/OCSVM/RF) — core model comparison.

### C. Interpretability and Feature Contributions

To assess interpretability, SHAP-based feature attribution was applied to analyze the contribution of input variables to anomaly detection outcomes. Results indicate that response time and temporal features, including hour-of-day and day-of-week, are the dominant predictors of anomalous behavior. This finding aligns with operational intuition regarding workload-driven delays and reinforces the transparency of the proposed framework for clinical monitoring and audit purposes.

**Fig. 5.**
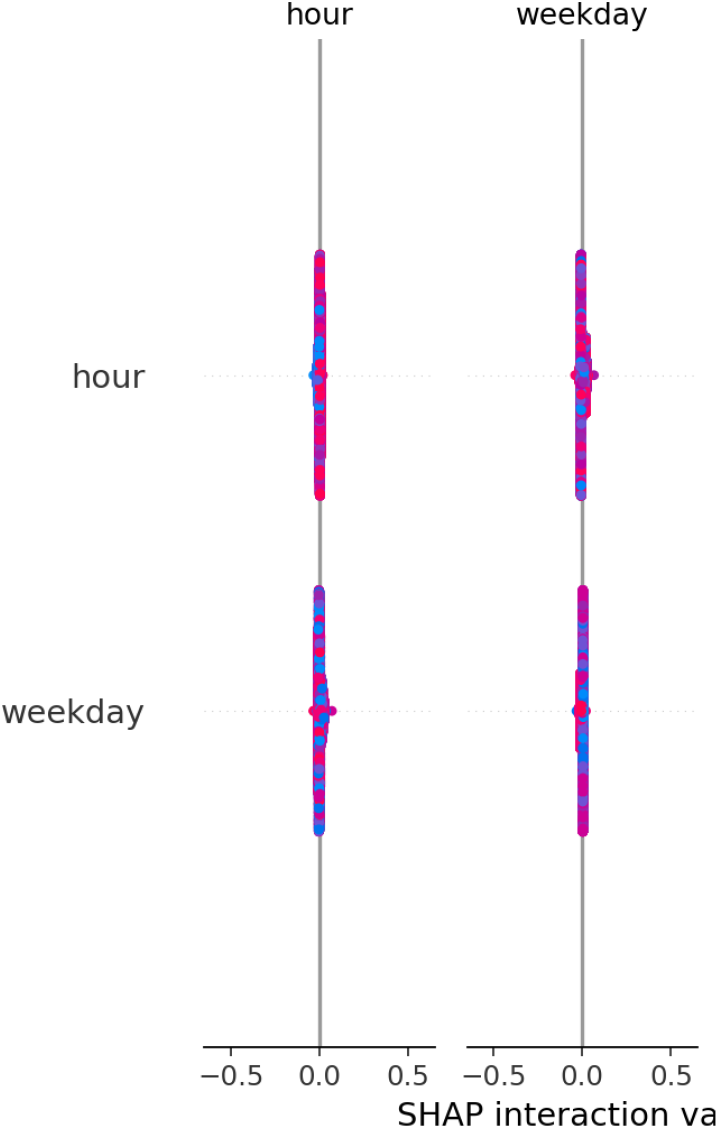
SHAP bar plot — explainability summary of feature contributions.

### D. Threshold Sensitivity and Operational Trade-offs

Threshold sensitivity analysis was conducted to identify operating points that balance detection performance and operational burden. By sweeping anomaly score thresholds, the analysis identified an operating region where F1-score is maximized (approximately at a threshold of -0.08), as shown in the threshold scanning results. This provides an evidence-based basis for configuring alarm sensitivity, enabling deployment decisions that account for both missed detections and alert fatigue.

**Fig. 6.**
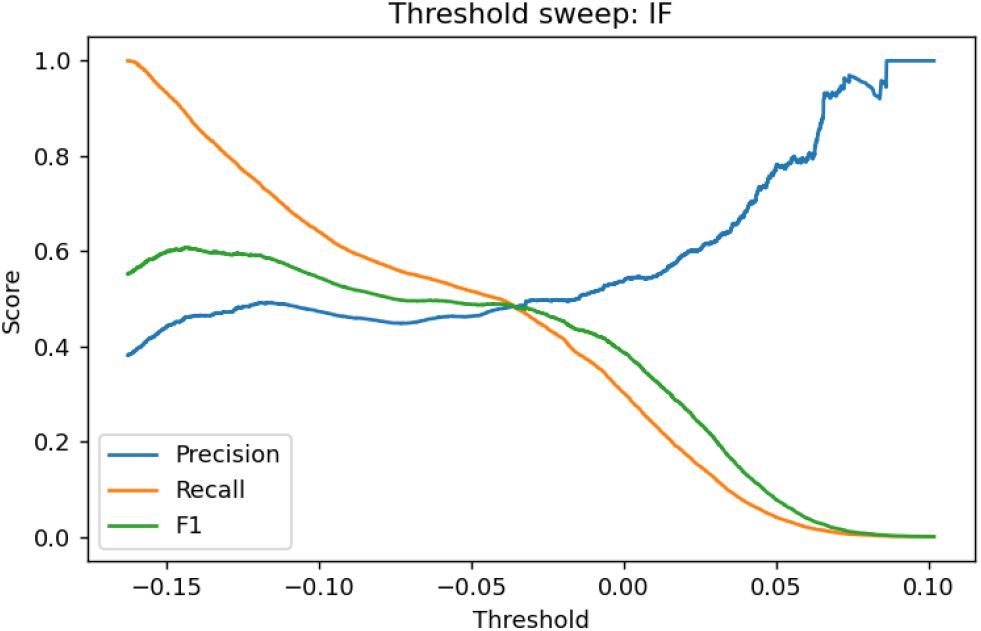
Threshold sweep (IF) — Precision/Recall/F1 sensitivity across thresholds.

### E. Additional Diagnostics

Additional diagnostic results, including autoencoder training convergence, confusion matrices, response-time distributions, and extended trade-off analyses, are provided in the appendix. These results further support the stability and reproducibility of the experimental findings without overloading the main narrative.

## VII. Conclusion and Future Work

“This work presented a lightweight runtime assurance framework for AI-enabled Nurse Call Systems, motivated by the need for continuous monitoring of safety-critical clinical infrastructure during live operation. By focusing on interpretable, computationally efficient techniques, the framework demonstrates that meaningful runtime assurance can be achieved without reliance on complex models or proprietary clinical data. Experimental results show that unsupervised detection, combined with operationally relevant evaluation metrics and threshold sensitivity analysis, can effectively surface safety-relevant deviations such as response delays and abnormal call patterns.

Beyond detection performance, the framework emphasizes transparency and operational relevance, supporting clinical trust and auditability. The use of interpretable diagnostics and evidence-based threshold tuning illustrates how anomaly detection can function as a practical assurance mechanism rather than as an isolated analytical tool. These characteristics are particularly important for deployment in resource-constrained and safety-critical healthcare environments.

Several limitations remain. The current evaluation relies primarily on simulated logs derived from public service request data, which may not fully capture the variability, noise, and contextual complexity of real-world nurse call operations. In addition, the scope of features is intentionally limited to support deployability, which may constrain detection performance in more complex settings.

Future work will focus on extending the framework along three directions. First, incorporating additional temporal and contextual features, such as sliding-window call frequency, historical delay patterns, and unit-level workload indicators, may improve sensitivity to evolving operational conditions. Second, deeper integration of explainability techniques, including expanded SHAP-based analyses, could further support auditability and clinical interpretation of detected anomalies. Finally, validation using real-world nurse call data, in collaboration with healthcare partners, will be essential to assess robustness and generalizability under live deployment conditions.

## Data Availability

All source code, cleaned datasets, experiment scripts, and the interactive Streamlit application used in this study are publicly available as open research artifacts. The archived release is available via Zenodo (DOI: 10.5281/zenodo.17767143). The source code repository and demo application are also publicly accessible.

https://doi.org/10.5281/zenodo.17767143

https://github.com/maxineliu2020/ai-nursecall-runtime-anomaly-detection

https://nursecall-demo.streamlit.app/

https://data.cityofnewyork.us/Social-Services/311-Service-Requests-from-2020-to-Present/erm2-nwe9/about_data

## Acknowledgment

The author gratefully acknowledges the guidance and support of Instructor David R. Concepcion during the 695.715 Assured Autonomy course at Johns Hopkins University. His feedback and encouragement were instrumental in shaping both the technical direction and clarity of this work. AI-assisted writing tools were used for language refinement and formatting support.

## AppendixA

### Reproducibility

To ensure transparency and reproducibility, all source code, cleaned datasets, experiment scripts, and the interactive Streamlit application used in this study have been released as open research artifacts. These resources enable independent reproduction of the experiments, exploration of runtime anomaly detection behavior, and evaluation using custom datasets.

The materials are available through a public repository and an archived release, as detailed below:

- Source code repository: https://github.com/maxineliu2020/ai-nursecall-runtime-anomaly-detection
- Zenodo archival DOI: https://doi.org/10.5281/zenodo.17767143
- Interactive demo: https://nursecall-demo.streamlit.app/

These materials enable independent reproduction of the experiments, inspection of runtime behavior, and validation of the reported results.

## Appendix B

### Additional Diagnostics

A. *Confusion Matrix (IF)*
B. *Response-Time Distribution*
C. *Category Imbalance and Cross-Agency Differences*
D. *Operational Trade-off Curves*
E. *PR Curves with AE (Unsupervised Extension)*

**Fig. 7.**
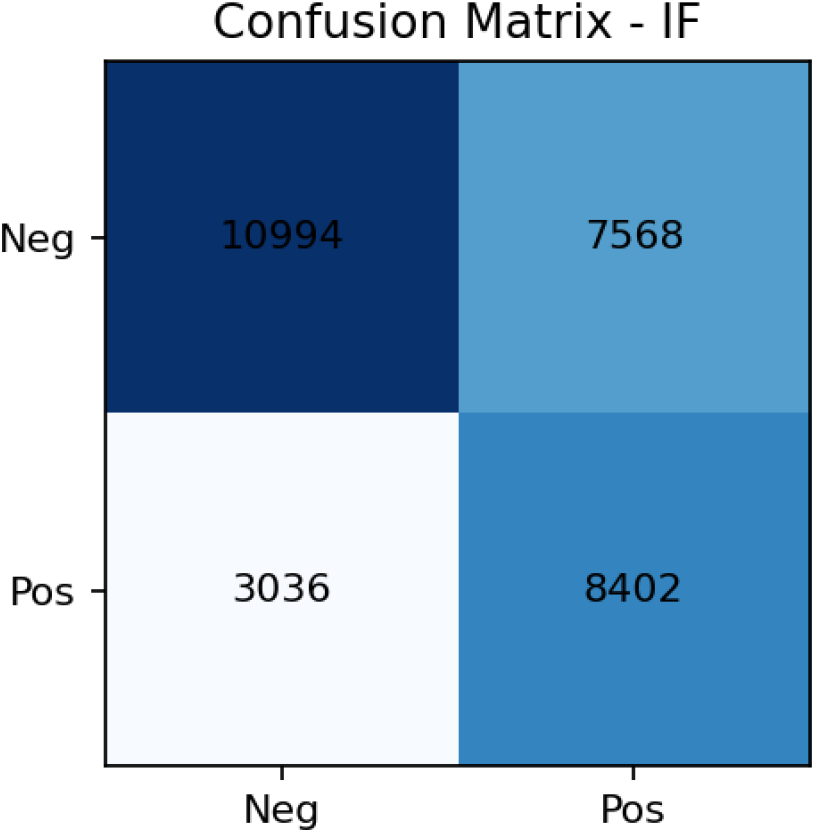
Confusion matrix (IF) — diagnostic analysis.

**Fig. 8.**
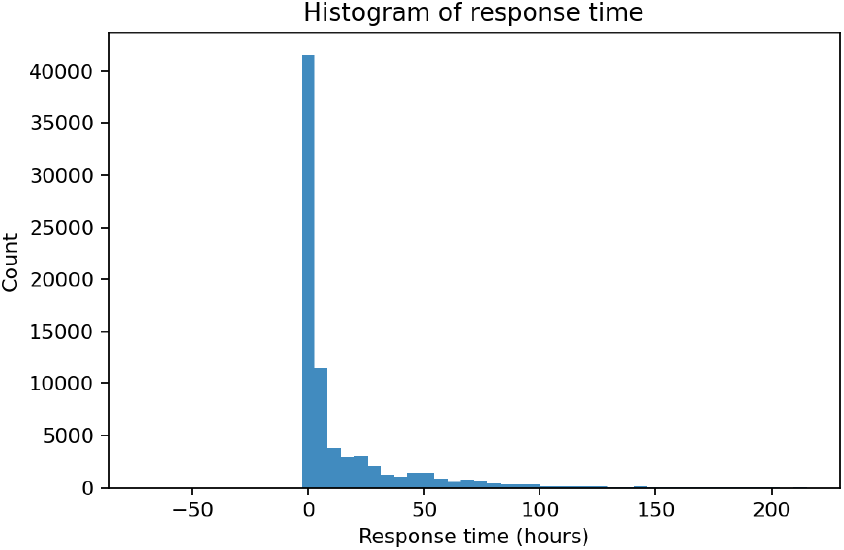
Histogram of response time — complementary distribution view.

**Fig. 9.**
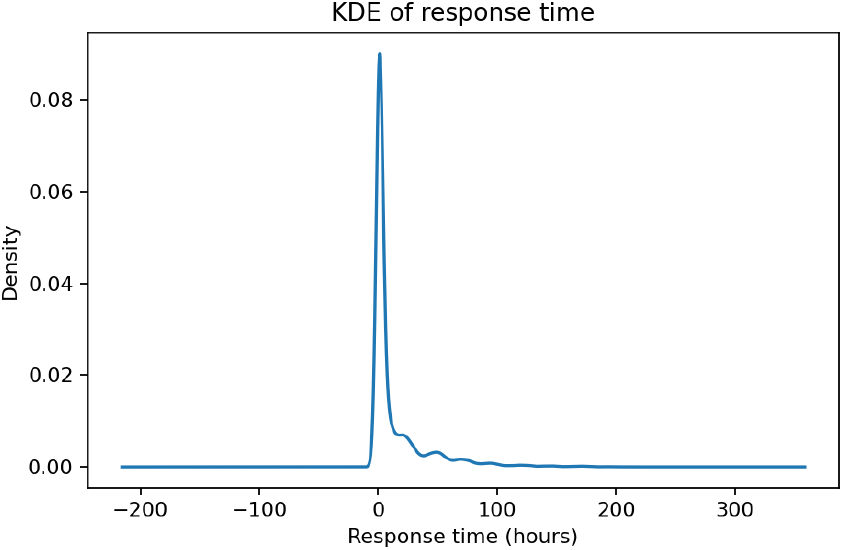
KDE of response time — smoothed statistical representation.

**Fig. 10.**
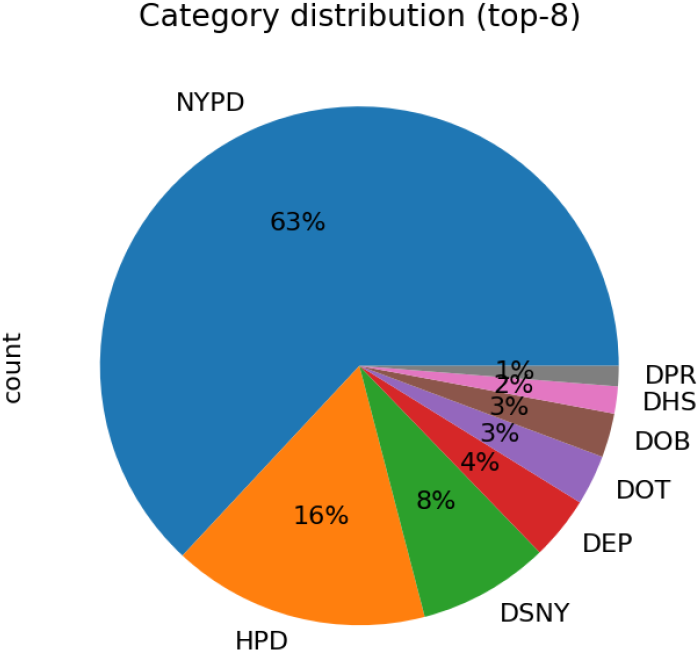
Category distribution (top-8) — class imbalance evidence.

**Fig. 11.**
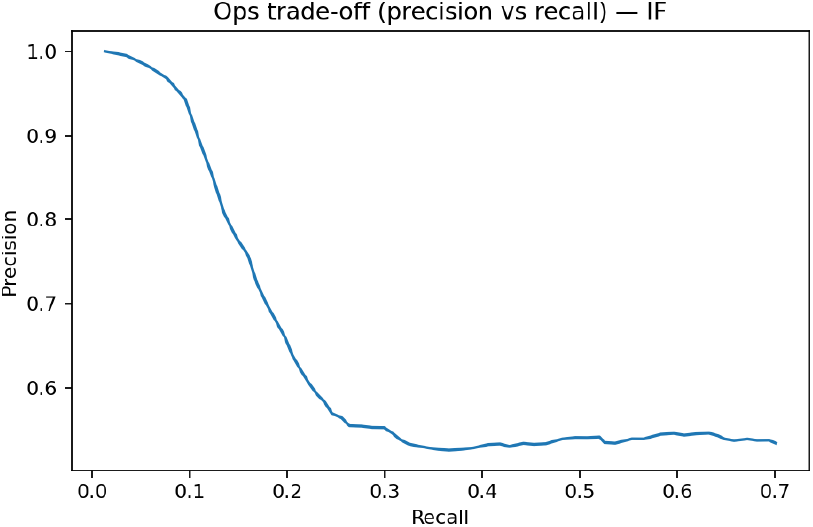
Ops trade-off (precision vs recall) — IF.

**Fig. 12.**
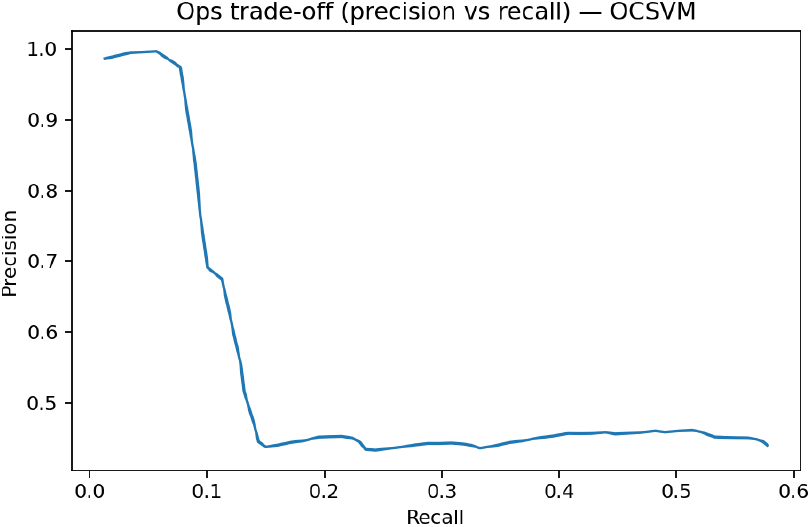
Ops trade-off (precision vs recall) — OCSVM.

**Fig. 13.**
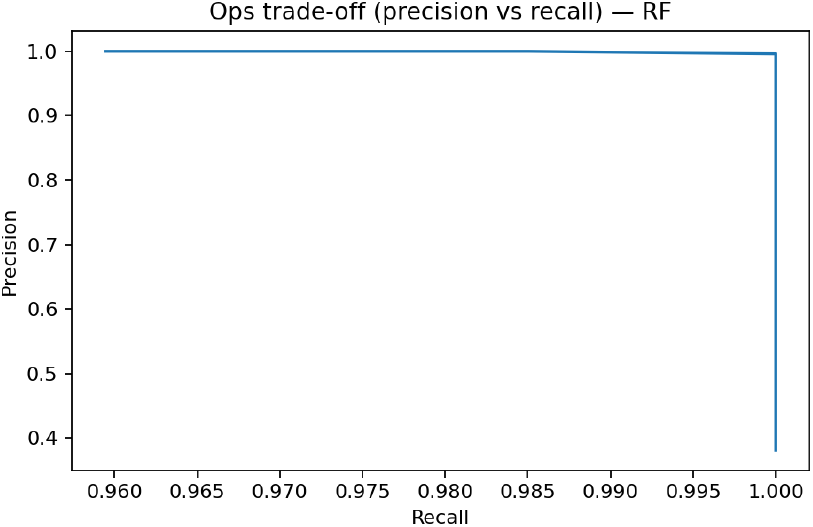
Ops trade-off (precision vs recall) — RF.

**Fig. 14.**
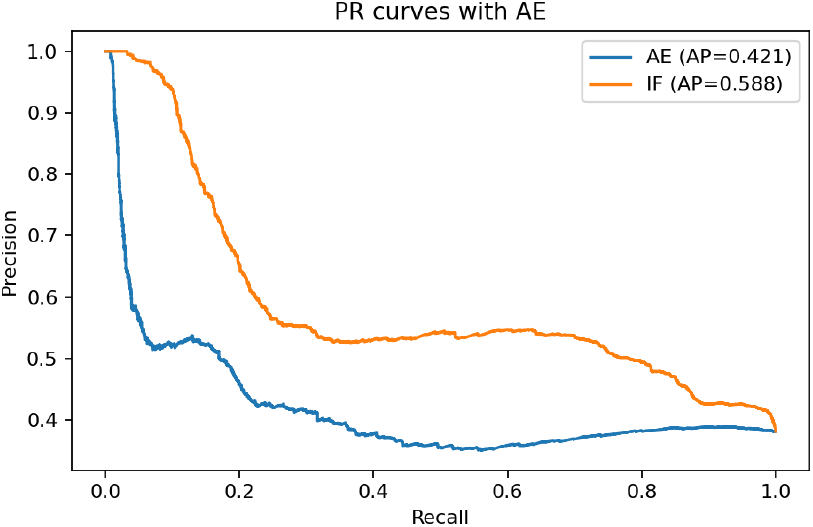
PR curves with AE — unsupervised extension (AE vs IF).

## References

[1] A. M. Said, A. O. Salami, and A. Alausa, “Efficient anomaly detection for smart hospital IoT systems,” Sensors, vol. 21, no. 4, p. 1424, 2021. [Online]. Available: https://www.mdpi.com/1424-8220/21/4/1424

[2] M. Tabassum, S. Mahmood, A. Bukhari, B. Alshemaimri, A. Daud, and F. Khalique, “Anomaly-based threat detection in smart health using machine learning,” BMC Medical Informatics and Decision Making, vol. 24, p. 347, 2024. [Online]. Available: https://pmc.ncbi.nlm.nih.gov/articles/PMC11577804/

[3] C. Brunk, J. Bueso, J. Pierre, B. Scheunemann, and T. Wulf, “Anomaly detection within mission-critical call processing,” 2024, arXiv preprint. [Online]. Available: https://arxiv.org/abs/2408.15499

[4] J. Dahmen and D. Cook, “Indirectly-supervised anomaly detection of clinically relevant events,” Frontiers in Artificial Intelligence, vol. 4, p. 648826, 2021. [Online]. Available: https://www.frontiersin.org/articles/10.3389/frai.2021.648826

[5] J. Aramann, A. Blasimme, E. Vayena, and A. Frey, “Explainability for artificial intelligence in healthcare: A multidisciplinary perspective,” BMC Medical Informatics and Decision Making, vol. 20, p. 310, 2020. [Online]. Available: https://bmcmedinformdecismak.biomedcentral.com/articles/10.1186/s12911-020-01332-3

[6] Z. Sadeghi, M. Rehman, A. Kausar, and S. Kumar, “A review of explainable artificial intelligence in healthcare,” Computers & Electrical Engineering, vol. 118, p. 109370, 2024. [Online]. Available: https://www.sciencedirect.com/science/article/pii/S0045790624002982

[7] F. Di Martino and F. Delmastro, “Explainable AI for clinical and remote health: Tabular and time-series data,” 2022, arXiv preprint. [Online]. Available: https://arxiv.org/abs/2209.06528

[8] Y. Jia, S. Wang, D. Chen, and C. Cheng, “The role of explainability in assuring machine learning safety in healthcare,” 2021, arXiv preprint. [Online]. Available: https://arxiv.org/abs/2109.00520

[9] J. Y. Kwon, M. J. Oh, J. H. Lee, T. H. Kim, Y. H. Kim, H. J. Jeon, J. W. Kim, J. H. Lee, J. H. Kim, H. J. Park, and S. H. Yoo, “Establishing organizational AI governance in healthcare,” NPJ Digital Medicine, vol. 8, p. 252, 2025. [Online]. Available: https://www.nature.com/articles/s41746-025-01909-3

[10] Duke Health Policy Center, “AI governance in healthcare systems: Aligning innovation, accountability, and trust,” Report, 2024, accessed 2025-12-29. [Online]. Available: https://healthpolicy.duke.edu/publications/ai-governance-healthcare-systems-aligning-innovation-accountability-and-trust

[11] W. Ma, “AI-driven cloud-security-framework for SME-healthcare (release v1.0),” Zenodo, 2025. [Online]. Available: 10.5281/zenodo.17605068

[12] Y. Liu, “AI nurse-call runtime anomaly detection (initial release),” Zenodo, 2025. [Online]. Available: 10.5281/zenodo.17767143

